# Analysis of host immunological response of adenovirus-based COVID-19 vaccines

**DOI:** 10.1101/2021.06.24.21259460

**Authors:** Suzan Farhang-Sardroodi, Chapin Korosec, Samaneh Gholami, Morgan Craig, Iain R Moyles, Mohammad Sajjad Ghaemi, Hsu Kiang Ooi, Jane M Heffernan

## Abstract

During the SARS-CoV-2 global pandemic, several vaccines, including mRNA and adenovirus vector approaches, have received emergency or full approval. However, supply chain logistics have hampered global vaccine delivery, which is impacting mass vaccination strategies. Recent studies have identified different strategies for vaccine dose administration so that supply constraints issues are diminished. These include increasing the time between consecutive doses in a two-dose vaccine regimen and reducing the dosage of the second dose. We consider both of these strategies in a mathematical modeling study of a non-replicating viral vector adenovirus vaccine in this work. We investigate the impact of different prime-boost strategies by quantifying their effects on immunological outcomes based on simple ordinary differential equations. The boost dose is administered either at a standard dose (SD) of 1000 or at a low dose (LD) of 500 or 250 vaccine particles. Simulated Second dose fractionation highlights previously shown dose-dependent features of the immune mechanism. In agreement with clinical characteristics of 175 COVID-19 recovered patients, the model predictions for either SD/SD or SD/LD regimens mainly show that by stretching the prime-boost interval until 18 or 20 weeks, the minimum promoted antibody (Nab) response is comparable with the neutralizing antibody level of COVID-19 recovered patients. The minimum stimulated antibody in SD/SD regimen is identical with the high level of clinical trial data. It is at the same range of the medium-high level of Nab in SD/LD, where the second dose is half or quarter of the standard dose.

## 1 Introduction

The spread of coronavirus disease-2019 (COVID-19), caused by severe acute respiratory syndrome coronavirus-2 (SARS-CoV-2), can be mitigated through safe and effective vaccines. Different vaccine types are currently being used to protect individuals from SARS-CoV-2 infection and disease. The four main types of COVID-19 vaccine in clinical trial includes whole virus, protein subunit, Viral Vector, and nucleic acid (RNA and DNA).

Limitations in vaccine supply, however, can affect the outcomes of the global vaccination campaign. Reductions in dose size, and a second dose delay whereby the second dose is delivered in a time frame beyond the manufacturer’s recommended schedule, can thus be considered, so that vaccine supply issues are diminished. In this work, we mathematically model adenovirus-based vaccines using a system of simple ordinary differential equations. The goals of our mathematical modeling study are two-fold, to (1) identify biological and vaccine characteristics that may allow for heightened and longer-lasting immune responses from vaccination, and (2) study the outcomes of a delayed second dose with the same or smaller dose size. The goals of this study are directly related to vaccine supply as we can determine if delaying and administering smaller second doses can provide immunological protection of similar magnitude to the recommended vaccine schedule (i.e., two similar-sized doses separated by 28 days).

The ODE-based model introduced in this work is based on the biological signaling pathway of the immune response to vaccination. The model includes both cellular and humoral immune system components, including vaccine particles, T helper cells, interferon-gamma (IFN*γ*), interleukin 6 (IL6), plasma B-cells, antibody, and cytotoxic T-cells. Model parameters were fit to clinical trial data for the COVID-19 ChAdOx1-S (AZD1222) vaccine developed by the University of Oxford and Astra-Zeneca. [1].

Our model findings show that by limiting the booster dose, we encounter decreased antibody and cytotoxic T-cell levels in the human body when compared to vaccine outcomes that follow the manufacturer’s recommended dose size and dosing schedule. Our results show evidence for a dose-dependent behavior of the immune system in response to an adenovirus-based vaccine [2]. The reduction in antibody level is more apparent if we consider long delays between the prime and second doses. This happens because the prime-boost time interval leads to a weak stimulation of the immune response. Nevertheless, in line with the previous clinical data [3], the model-predicted antibody level is comparable to the level attained by a patient with mild symptoms who recovers from COVID-19. Model predictions of attenuated IFN*γ* level by second shot delay and reduced dose size show the safety of a vaccine program with a delayed second dose.

This paper is organized as follows: In section 2, we describe the mathematical model of the adaptive immune response. We then fit the model to available clinical trial data for the Oxford/Astrazenca vaccine [1]. A sensitivity analysis is then performed to determine model parameters that most affect peak values of the immune system outcomes from vaccination, and thus, the longevity of components of vaccine-induced immunity. Finally, we study the effects of delays in the second dose of the vaccine and the use of smaller-sized second doses. We conclude the paper in section 4.

## 2 Model

The adaptive immune system is activated after exposure to an antigen either through vaccination or infection by a pathogen if the innate immune response is insufficient to stop the disease. Cell-mediated immunity, contributed by T-cells, and humoral immunity, controlled by activated B-cells, are components of the adaptive immune response that activate the immune system to protect the human body. These immune response components also generate memory T- and B-cells to protect an individual from future infection or disease. We have developed a mathematical model of an adenovirus vaccine that considers humoral and cell-mediated immune response mechanisms. The mechanisms of the cell-mediated immune response are illustrated in Figure 1, using bold and shaded components. Upon vaccination, vaccine particles will be recognized by components of the innate immune response, antigen-presenting cells (APCs, denoted here by APC1 and APC2, which are related to major histocompatibility complex (MHC) class 1 and 2 molecules, respectively). T helper type 0 cells (Th_0_) are activated through (APC2) and differentiate into Th_1_ and Th_2_ cells (central part). Cytotoxic T-cells (CTL, also called CD8 T-cells) can then be stimulated through the cytokine production from Th_1_ cells, including interleukins (IL2, IL12), transforming growth factor-alpha TGF*α* and Interferon IFN*γ*. Activated CTL differentiates into effector cells which can then become memory CD8 T-cells. Th_2_ cells recognize the Th epitopes that are presented by B-cells through the MHC class II receptors. After being activated, Th_2_ cells secrete IL4, IL5, IL6, IL10, and TGF*β* to stimulate B-cell activation and differentiation into plasma cells and memory B-cells (right part). The plasma cell produces neutralizing (NAb) antibody responsible for clearing the infection [4–6].

**Figure 1:**
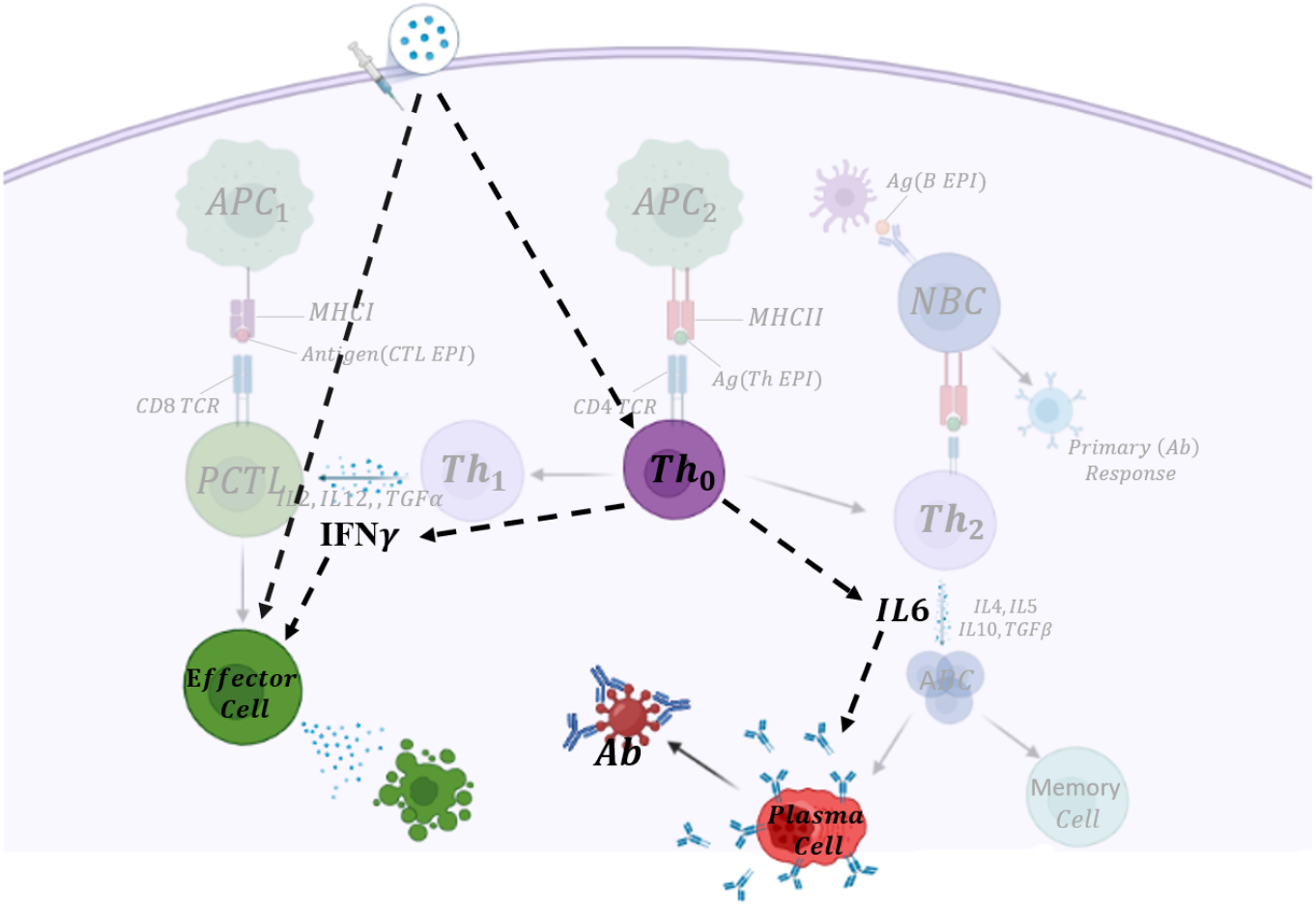
Vaccine-induced immune activation pathway for an adenovirus vaccine. Faint background: The subsequent downstream of signaling pathways activated through adaptive immunity when SARS-CoV-2 enters the human cell. Highlighted compartments describe vector-based vaccine-induced immune system stimulation that is modeled explicitly in this study. The dashed arrows show implicit communications between cells and cytokines, and the only solid arrow indicates the production of antibodies by B-cells.

A simple network that reflects the entire diagram in Figure 1 is shown in bold. This simple network provides the basis of the mathematical model used in this study and has been chosen so to reduce the dimension of the system, given available parameters from the literature and the availability of the vaccine data. We explicitly consider T helper type 0 cells, plasma B-cells, antibody, cytotoxic T-cells, and two central cytokines, including IFN*γ* and IL6. An external stimulus, the vaccine, activates the immune response. The model consists of the following system of seven nonlinear ordinary differential equations.

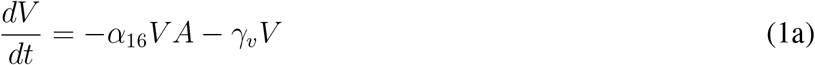

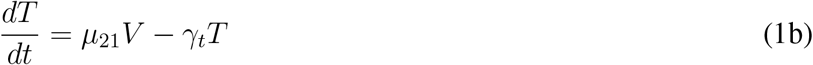

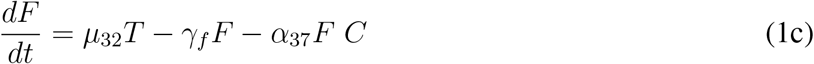

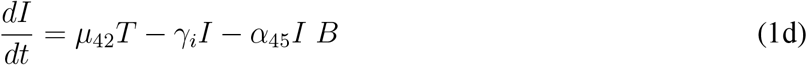

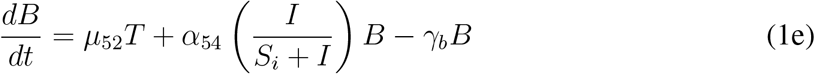

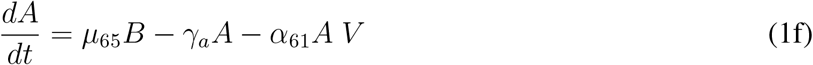

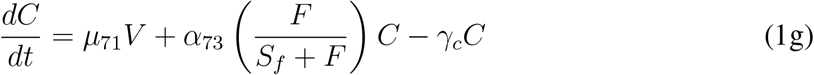

with variables summarized in Table 1, and 21 model parameters, listed in Table 2. Accordingly, parameters referring to variable production processes are denoted by *µ*_*ij*_, where *i* and *j* = {1, 2, …, 7}, corresponding to populations {*V, T, F, I, B, A, C*}, and denote the stimulated and stimulating populations, respectively. Parameters referring to the interaction of entities *i* and *j* are denoted by *α*_*ij*_. We note that *α*_*i,j*_ is not necessarily equal to *α*_*ji*_. Finally, parameters referring to the natural death of the population under consideration are defined by *γ*_*k*_, where *k* in {*v, t, f, i, b, a, c*}, corresponding to the model variables.

**Table 1:** Model Variables.

| Variable | Definition |
| --- | --- |
| $V$ | Vaccine cell |
| $T$ | T helper type 0 cell ($Th_0$) |
| $F$ | Interferon gamma ($IFN\gamma$) |
| $I$ | Interleukin 6 ($IL - 6$) |
| $B$ | Plasma B-cell |
| $A$ | Antibody |
| $C$ | Cytotoxic T-cell |

**Table 2:** Model Parameters (with unit: (*day*)^*−*1^) for immune response to COVID-19 vaccination by fitting to clinical 18-55 (SD) age group of IFN*γ* data, RMSE≈ 55, and ≥70 (two-dose) age group of antibody data, RMSE≈ 3451. Parameter values determined by curve fitting the model solution to clinical trial data from [1].

| Parameter | Definition | Value | Comment |
| --- | --- | --- | --- |
| $\alpha_{16}$ | Vaccine and antibody binding rate of inactivation | 1E-6 | Handle et al. 2018 |
| $\gamma_v$ | Vaccine clearance rate | 0.2 | Cao et al.2016 |
| $\mu_{21}$ | $Th_0$ cells activation rate by vaccine particles | 0.035 | Chosen |
| $\gamma_t$ | $Th_0$ cells natural death rate | 0.055 | Cao et al.2016 |
| $\mu_{32}$ | $IFN\gamma$ stimulation rate by $Th_0$ | 2.55 | Fitted |
| $\gamma_f$ | $IFN\gamma$ natural degradation rate | 0.13 | Fitted |
| $\alpha_{37}$ | $IFN\gamma$ absorption rate by CTL for mitotic signals | 0.006 | Fitted |
| $\mu_{42}$ | IL6 release rate by $Th_0$ | 1.3 | Fitted |
| $\gamma_i$ | IL6 natural degradation rate | 0.0008 | Chosen |
| $\alpha_{45}$ | IL6 absorption rate by B-cells for mitotic signals | 0.0001 | Fitted |
| $\mu_{52}$ | B-cell activation rate by $Th_0$ | 0.02 | Fitted |
| $\alpha_{54}$ | B-cell stimulation rate by IL | 0.05 | Fitted |
| $S_i$ | B-cell duplication threshold due to IL | 1000 | Chosen |
| $\gamma_b$ | B-cell natural death rate | 0.06 | Fitted |
| $\varepsilon\mu_{65}$ | Released Ab rate by B-cells | 7 | Fitted |
| $\gamma_a$ | Ab natural degradation rate | 0.06 | Fitted |
| $\alpha_{61}$ | Ab - V cells binding rate | 1E-7 | Chosen |
| $\mu_{71}$ | CTL activation rate by vaccine | 0.002 | Fitted |
| $\alpha_{73}$ | CTL stimulation rate by $IFN\gamma$ | 0.09 | Fitted |
| $S_f$ | CTL duplication threshold due to $IFN\gamma$ | 600 | Chosen |
| $\gamma_c$ | CTL natural death rate | 0.01 | Wang et al. |

In Eqn (1a), vaccine particles are injected into the host with a predefined dosage. Their inhibition is described by natural decay and neutralization by antibody [7, 8].

T helper cells are essential cells in that they are involved in activating the humoral and cell-mediated immune responses. T helper cells are activated by specialized antigen-presenting cells (APCs) through the primary histocompatibility class II/peptide complexes. Model (1) considers T helper type 0 cells only (we do not consider differentiated T-cell dynamics). See Eqn (1b). We also simplify the model by ignoring the explicit population of APCs and instead assume that the Th_0_ population activation is proportional to the vaccine particles in the system (i.e., we assume that the APC population is proportional to the vaccine particles count).

IFN*γ* is a type-II IFN that plays a crucial role in regulating the adaptive immune response. It is produced by a wide variety of lymphocytes, including CD4+, CD8+, and regulatory T (Treg) cells, B-cells, and NK cells. Although numerous cells can express IFN*γ*, it is mainly secreted by T-cells and it is the defining cytokine of Th_1_ helper cells [9, 10]. Accordingly, and based on the signaling pathway introduced by Figure 1 here, in Eqn (1c), we define the source term of IFN*γ* (*µ*_32_*T*) to be the Th_0_ cells in the system. We also assume that IFN*γ* can be degraded or removed in the system by (1) Th_0_ cell surface binding (for mitotic and stimulation signals in cytotoxic T-cell, *C*, proliferation [11]), and (2) natural decay.

We also explicity model IL6, another cytokine in the system. See Eqn (1d). IL6 is a pleiotropic cytokine[12, 13] produced by many different T-cell types, including T- and B-cells [14], that has a pivotal role in the activation and stimulation of the immune response [15]. IL6 also plays an important role in antibody production [16]. IL6 has a wide range of functions, and acts as a B-cell stimulatory factor to induce antibody production [17]. Elevated IL6 levels found in COVID-19 patients with mild and severe symptoms [18–31] concluded that IL6, alongside other cytokines, can be of prognostic value in these patients [32]. In our mathematical model, IL6 is considered to be secreted indirectly by Th_0_ cells and is partially absorbed for stimulation signals in B-cell priming.

Plasma B-cells are long-lived, non-proliferating cells arising from B-cell differentiation, stimulated by interaction with T helper cells. Activated plasma B-cells produce neutralizing antibody, which are responsible for clearing the infection. In Eqn (1e) we consider an indirect activation of plasma B-cells by Th_0_ cells at rate *µ*_52_*T*, and by IL6, which is assumed to have an adjuvanted role in stimulation, 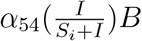, where *α*_54_ is a recruitment rate and *S*_*i*_ is a saturation constant. Plasma B-cells die naturally at rate *γ*_*b*_.

Humoral immunity is an antibody-mediated response that occurs when plasma B-cells are activated. In Eqn (1f) We assume that antibody production is proportional to the number of plasma B-cells, by rate *µ*_65_. We also assume that antibody are degraded at a rate *γ*_*a*_, or removed through vaccine binding at rate *α*_61_. they can be lost to the system through vaccine particle binding (*α*_61_*A V*). Note that the simulated antibody without specialization, is entirely the neutralizing antibody (Nabs) which is responsible for defending cells from pathogens or infectious particle by neutralizing its biological effects.

Finally, like the activation of the Th_0_ cells, in Eqn (1g), we assume that cytotoxic T-cells (also known as cytotoxic T-lymphocytes, CTL, and activated CD8 T-cells) priming is proportional to the number of vaccine cell particles (*µ*_71_*V*). We also assume that IFN*γ* Can stimulate further cytotoxic effector T-cells (term 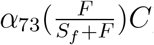), [9, 33]. Finally, cytotoxic T-cells die at rate *γ*_*c*_. The initial conditions for all activated cells and cytokines are zero *T* (0) = *F* (0) = *I*(0) = *B*(0) = *A*(0) = *C*(0) = 0, assuming that we are starting in a system with no activated immune response. The first and second dose are entered into the system using an initial condition of *V* (0) = 1000, and an impulse of 1000, 500 or 250 vaccine particles provided to the system at the time of the second dose. The standard dose size used here is chosen to be 1000 vaccine particles. This is an arbitrary value. When a smaller or larger dose size is chosen, parameters *µ*_2_1, *α*_61_, and *µ*_71_ are simply rescaled.

### Parameter fitting

The vaccine on which we base our parameters is the one produced by AstraZeneca/Oxford [1], an adenovirus-based SARS CoV-2 vaccine which has been approved in countries United Kingdom, Bangladesh, United States, Egypt, Japan, Australia, Canada, Switzerland, Thailand, Malaysia, Philippines, South Korea and South Africa. We parameterized the model by fixing some parameter values from the literature and others to the vaccine trial data using a grid search method. Some parameters had limited data availability and their values were chosen (see Table 2). Sensitivity analyses were performed to assess variations in model outcomes due to changes in the fixed and chosen parameters.

The final fit of the model to the vaccine trial data was defined by the minimum of the root-mean-squared-error (RMSE), which considers differences between the IFN*γ* and antibody data for SARS-CoV-2 IgG response, and the model-predicted values for these values. Clinical trial data for one- and two-doses of the vaccine were both considered.

We note that the parameters of the modelled neutralizing antibody response are fit to IgG clinical data. Although IgG is the most common antibody (70 − 75% of all human immunoglobulins found in the plasma [34, 35]), we must consider a proportionality between IgG and the antibody population. We thus assumed that IgG = *ε* NAbs, and the antibody equation in Model (1) becomes

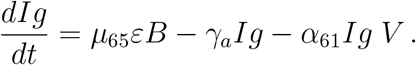

Consequently, we consider *µ*_65_*ε* as one parameter in the model fit to the vaccine trial data.

#### 2.1 Sensitivity Analysis

We employ sensitivity analysis methods that include Latin Hypercube Sampling (LHS) [36, 37] and Partial Rank Correlation Coefficients (PRCC) to study the effects of parameter variation on key model outcomes of interest. We use 10000 samples from a uniform distribution over parameter ranges within the interval with median=parameter value, listed in Table 2, minimum=0.5*parameter and maximum=1.5*parameter. PRCC values are calculated on model outcomes associated with a strong vaccine-induced immune response. We have chosen to study the peak magnitude of each model variable, as peak value correlates with a longer-lasting immune response. We note that PRCC values with a magnitude close to unity indicate that the parameter has the highest attainable significant impact on the model output [38]. A value greater than 0.5 is assumed to be significant [38]. Additionally, PRCC values can be negatively (negative sign) or positively correlated with a model outcome [39].

## 3 Results

The fitted model, to the IgG and IFN*γ* data [1], is shown in Figure 2. We observe that post priming the model-predicted antibody level is comparable to the clinical trial data for the 56-69 (one dose) age group, and it lies in the upper range of the antibody levels reported by [1]. After boosting (red line), the antibody level increases to about 10^4^ titer which lies close to the lower end of the clinical trial measurements.

**Figure 2:**
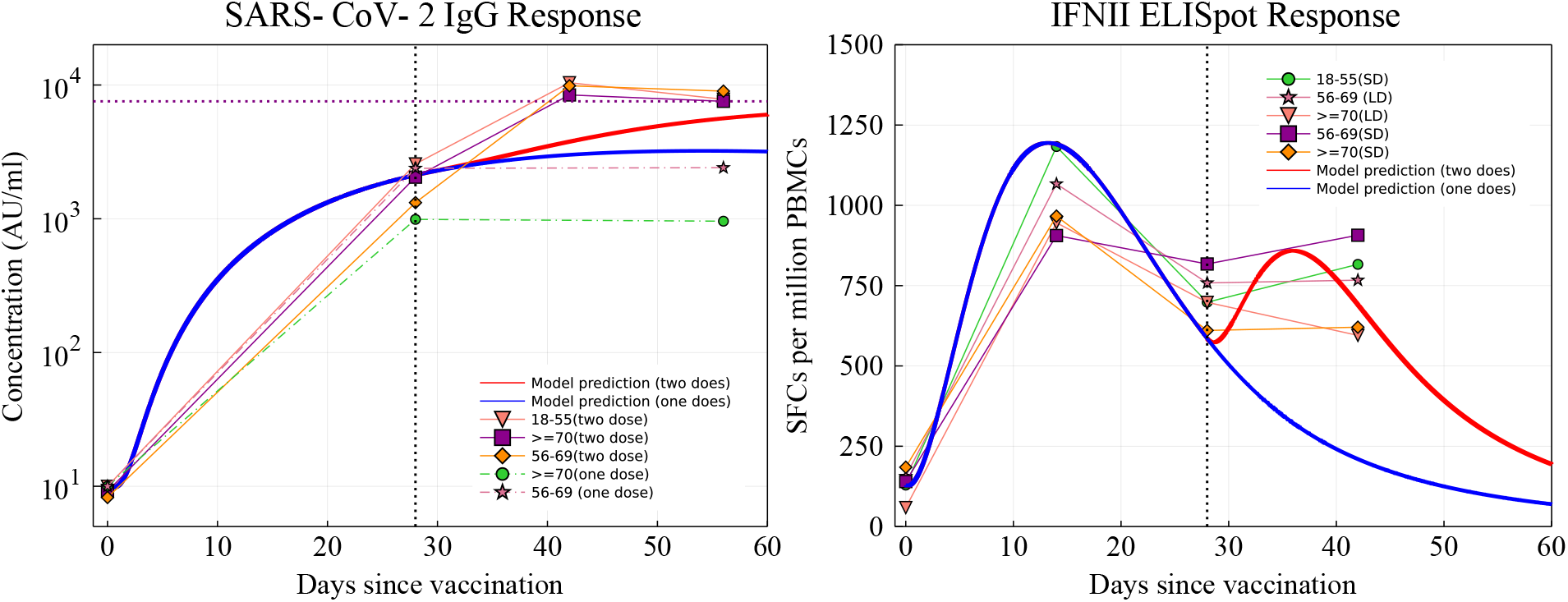
Antibody and IFN*γ* fit to the clinical trial data [1]. Blue and red solid lines: predicted results for participants who received one (blue) pr two doses (red), with a boost dose at day 28. Left: The purple dashed horizontal line shows the maximum stimulated antibody level post-boost.

The model predicted result of IFN*γ* after the prime is also consistent with clinical data and boosted by the second dose injection. The resulting IFN*γ* level predicted by the model lie close to the lower values measured in the clinical trial at days 28 and 42.

### 3.1 Sensitivity Analysis

A sensitivity analysis is performed to assess changes in model outcomes as parameter values are varied. We examine the sensitivity of the peak values of T helper cells, IFN*γ*, interleukin, B-cells, antibody, and cytotoxic T-cells. We are interested in determining what parameters maximize peak values so that longer-lasting immune outcomes can be realized from vaccination (assuming that longer-lasting immunity correlates with increased peak value). Results of the LHS/PRCC analysis are shown in Figure 3 for all model parameters. Significant parameters affecting peak values, with an absolute value of PRCC*>* 0.5, are listed in Table 3. A monotonic relationship between the outcomes and the parameter values is confirmed in all cases.

**Figure 3:**
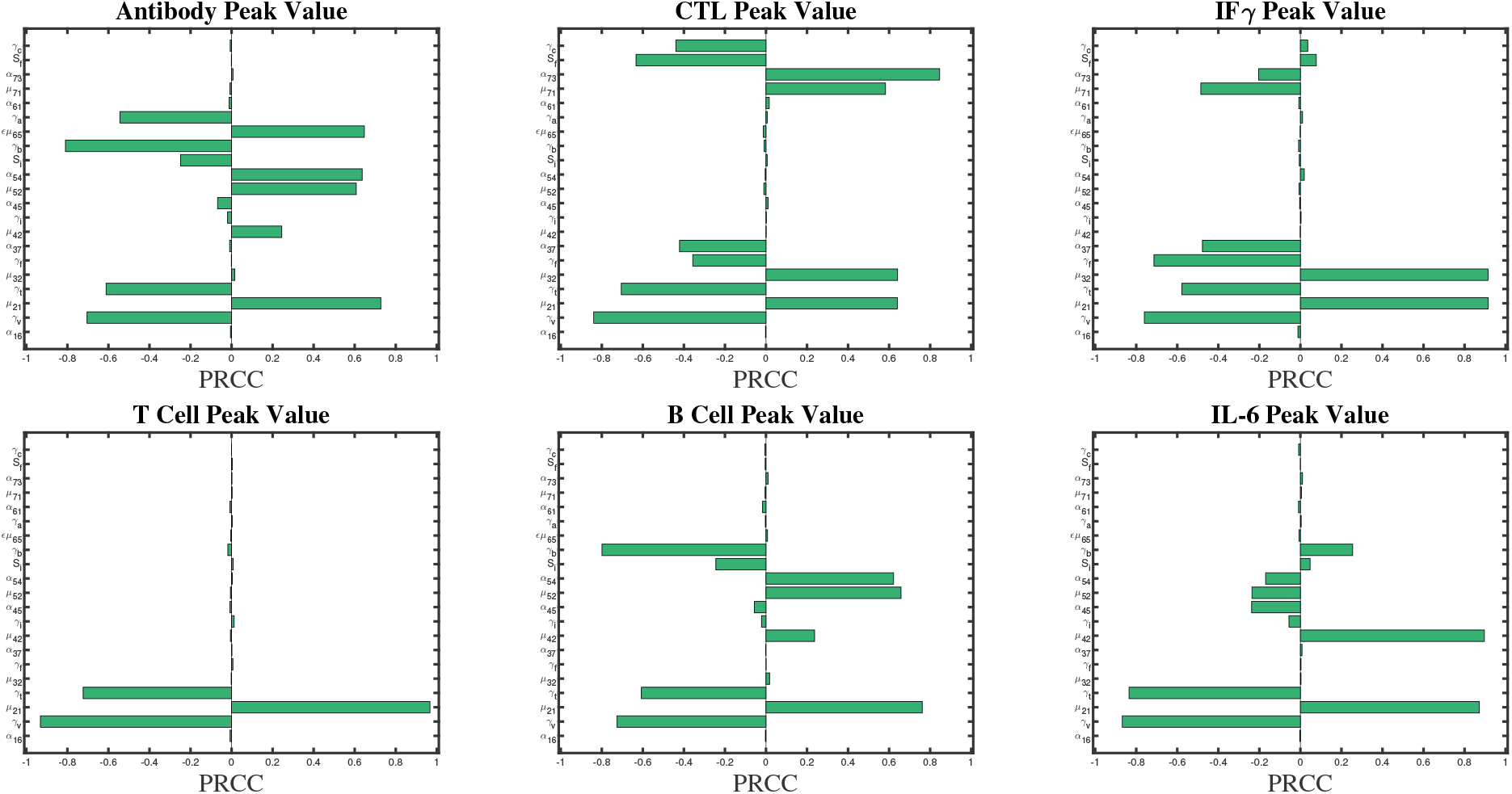
Sensitivity analysis of the Model (1) using 10000 iterations of a latin hypercube sampling (LHS) method with a partial rank correlation coefficient (PRCC). PRCC values with magnitude close to unity indicate that the parameter has a strong impact on the model output [38].

**Table 3:**
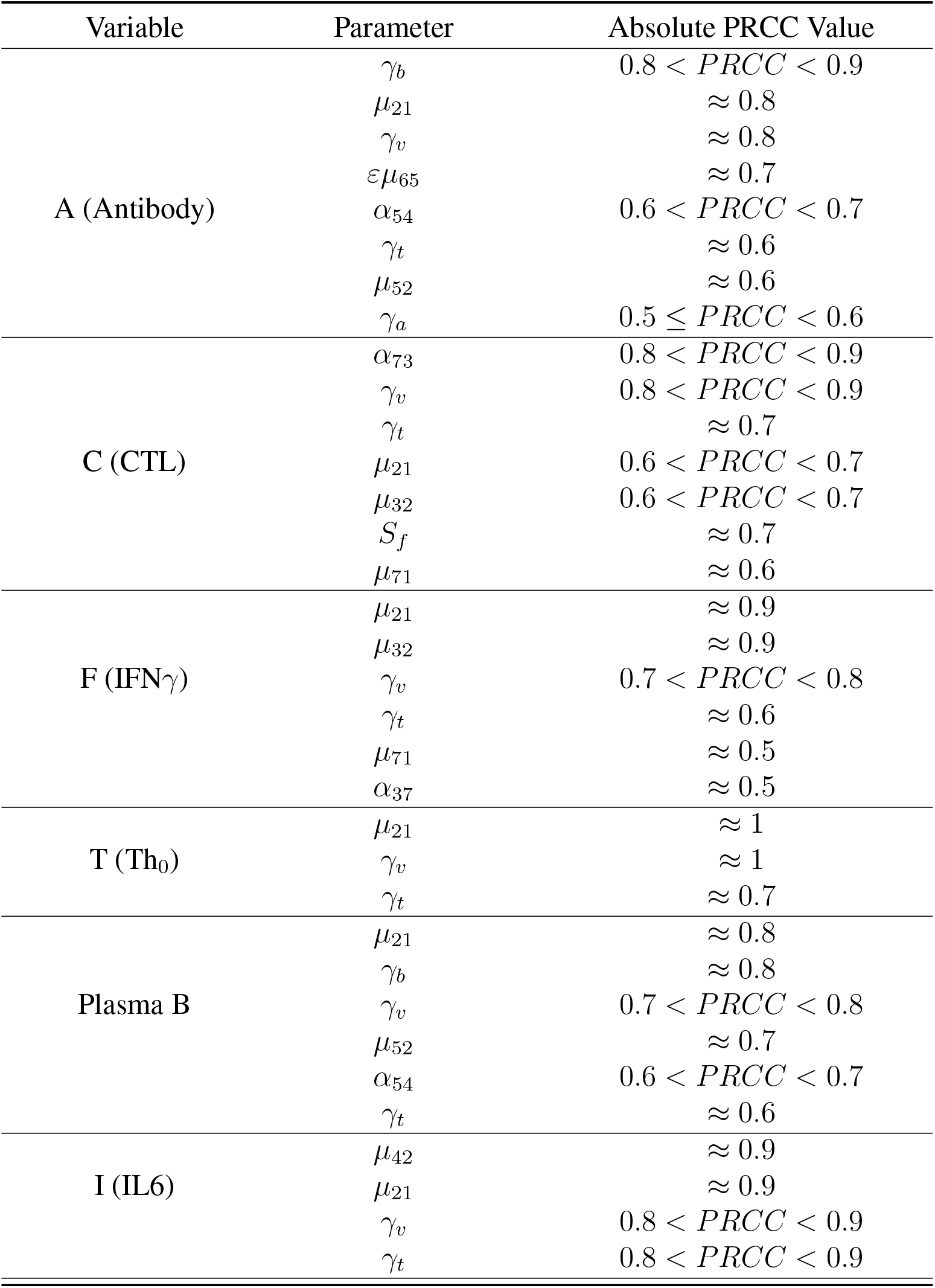
Parameter Sensitivity with absolute value of *PRCC ≥* 0.5.

Generally, we find that increases in peak value correlate with increases in stimulation and secretion, and decrease with increases in death and decay rates. Peak antibody and plasma B-cells both have a high sensitivity to *γ*_*b*_, the B-cell natural death rate, *α*_54_, the B-cell stimulation rate by *IL* − 6 and *µ*_52_, the B-cell activation rate by Th_0_. Peak CTL is very sensitive to *α*_73_, simulation rate by IFN*γ,µ*_32_, IFN*γ* stimulation rate by Th_0_, *S*_*f*_, CTL duplication threshold due to IFN*γ* which is a chosen parameter, and activation by virus particles, *µ*_71_. Variations in secretion rates *µ*_32_ and *µ*_42_ significantly affect the peak values of interferon and interleukin, respectively.

We note that variation in *µ*_21_, *γ*_*t*_ and *γ*_*v*_ significantly affects all populations. This is an intuitive result as *µ*_21_, *γ*_*t*_, and *γ*_*v*_ all affect the peak Th_0_ value, and the Th_0_ population stimulates the rest of the immune response.

We note that *µ*_21_ is a chosen parameter. Although variation in this parameter significantly affects all population peak values, since it is related to the activation rate of the Th_0_ population, which activates the rest of the immune response, it is always countered by sensitivity to *γ*_*v*_ and *γ*_*t*_, which are parameters informed by the literature. Given a constant activation and proliferation capacity of Th_0_ cells, an increase in *µ*_21_ would require an increase in *γ*_*t*_ of similar magnitude. Therefore, we conclude that sensitivity to this parameter is not a concern.

Lastly, we note that the only other chosen parameter value that significantly affects any modle outcome is *S*_*f*_, the saturation constant of the CTL population. The parameter ranks 6th in significance related to peak CTL value only. We therefore conclude that sensitivity to this parameter is not a concern.

### 3.2 Mechanism of vaccine-induced immunity with booster delay and sparing

We now apply Model (1) in a study of reduced second dose volume, and its delay. We consider several different scenarios based on varying assumptions on boosting (second dose). We first provide the system with a standard dose (SD) of 10^3^ vaccine particles. We then provide a SD second dose, a low dose (LD) of 500 vaccine particles, or a LD of 250 vaccine particles. The second dose is injected into the system 28, 42, 56, 70, 84, 98, 112, 126, or 140 days later (corresponding to 4, 6, 8, 10, 12, 14, 16, 18, and 20 weeks between doses). Figure 4 shows the antibody and CTL populations generated from the model considering all of these cases.

**Figure 4:**
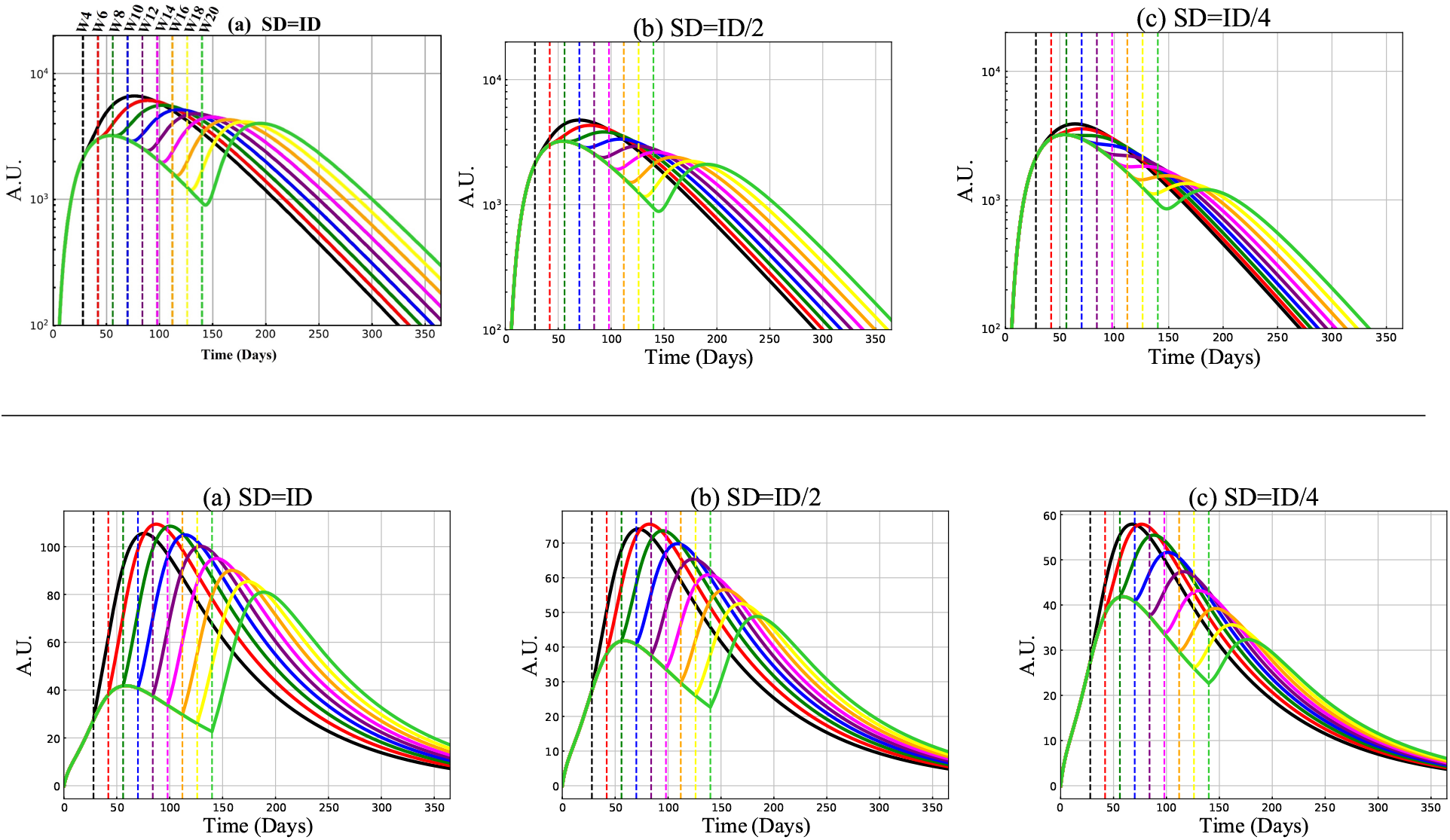
Antibody (IgG) and CTL outcomes with standard (SD) and low dose (LD), with and without delay. Model predictions of antibody, first row, and cytotoxic T-cells, second row, with second dose vaccination on days:28 (week:4),42 (week:6), 56 (week:8), 70 (week:10), 84 (week:12), 98 (week:14), 112 (week:16), 126 (week:18), and 140 (week:20). The second dose value (SD) in panels (a) is the same as the initial dose (ID) value:(SD=ID=1000 vaccine particles), in panels (b) it is decreased by half (SD=ID/2=500 vaccine particles), and in panels (c) is decreased by a quarter (SD=ID/4=250 particles).

#### Antibody and Cytotoxic T-cell Responses

Model predictions suggest quantitative differences in neutralizing antibody and cytotoxic T-cell responses stimulated by the vaccine administration by an SD or LD boost 4 to 20 weeks post priming, see Figure 4. Good agreement is observed with [2, 40] in that the immune responses following the adenovirus vaccination show dose-dependency. By Comparing the SD/SD and SD/LD regimens, we find that larger second doses leads to the more stimulated antibodies and cytotoxic T cells. We, however, also observe that a higher antibody or CTL peak after the second dose, compared to the first dose peak, may not be achievable if the doses are too far apart (long delays in second dose), or the dose is too small. For the antibody population, the peak after the second dose is always greater than the first dose peak if an SD is used. When an LD is used, the time between doses must be shorter. For the CTL, a higher second peak can be achieved under all scenarios using an SD and an LD=SD/2, but the peak after the second dose may not surpass the first peak if an LD=SD/4 is used (shorter times between doses are needed). Considering the CTL population we can also observe that shorter times between doses do not necessarily result in larger CTL peak values. Here, we observe that a time-frame of 4 weeks between doses is not optimal. Generally, considering the antibody population, we find that the second-dose induced enhancement is increased if the prime-boost time interval is short. The differing outcomes between the antibody and CTL populations may be explained by the fact that CTL activity might be required to account for lower levels of antibodies that cannot neutralize the virus particle efficiently.

#### Cytokines, B and Th_0_ Cell Responses

Here we investigate the model predictions of proinflammatory cytokines, including IFN*γ* and IL6, alongside plasma B- and Th_0_-cells for four different prime-boost intervals (containing 4, 6, 8, and 10 weeks) in SD/SD regimen. Results are shown in Figure 5. Here, we observe that prolonging the time interval between doses reduces levels of IFN*γ* but increases IL6 levels. We also observe that the plasma B-cell count increases, but that a Th_0_ enhancement can only be achieved if the time between doses is less than 70 days.

**Figure 5:**
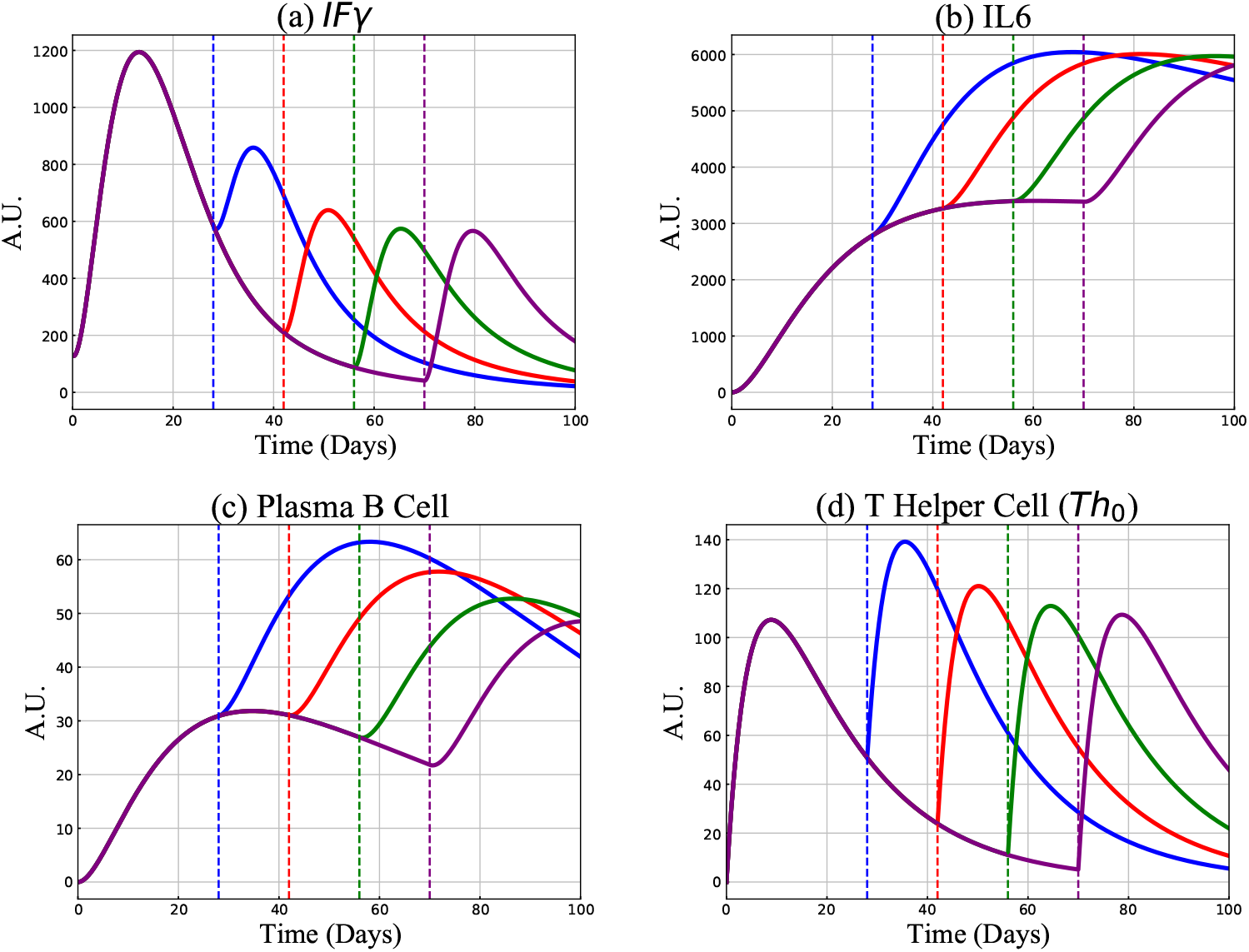
Model predictions of IFN*γ*, IL-6, plasma B-cells and T helper type 0 cells Th_0_ for the received boost dose (1000 vaccine particles) on days 28 (blue curve), 42 (red curve), 56 (green curve), 70 (purple curve). A vertical dashed line shows the second dose injection days.

#### Protective Capacity

An important question that we must consider is whether the model predicted antibody and CTL levels would protect against SARS CoV-2 infection by existing or new emerging variants. To answer this question, we compare model predictions to the clinical trial data in [3]. In this paper, the authors measured SARS-Cov-2-specific neutralizing antibody in plasma from 175 recovered COVID-19 patients with mild symptoms. They reported that 39% of the patients have medium-high antibody 1000-25000 titers (ID50), that 30% have less than 500 antibody titers, and 14% have titers greater than 2500. Figure 4 shows that after second dose injection for the SD/SD regimen the maximum stimulated IgG antibody for short prime-boost intervals such as 4, 6, 8, 10 weeks, is much higher than a 2500 titer (≈10^4^). The maximum stimulated antibody however decreases as the time between doses increases. At 20 weeks between doses, the antibody level reaches ≈ 4000 upon boosting. Interestingly, for the regimen SD/LD, with the second dose value at SD/2, the promoted antibody level up to an 18-week delay is greater than 2500 and is about the same level at 20 weeks between doses (*≈* 2500). For the last scenario where the second dose is a quarter of the prime dose, the minimum activated antibody level is ≈ 1000 for long time-frame between doses (week 18, or 20).

We must now consider neutralizing antibodies to determine protective capacity. Consider represented by the parameter *ε* (or something similar). From [35], if we assume *ε* = 0.7, we find that the minimum promoted neutralizing antibody through different second-dose injection weeks (from week=4,…. 20) for (i) SD/SD regimen is ≈ 5714, (ii) SD/LD regimen with LD=SD/2 is approximated by ≈ 3571, and (iii) SD/LD with LD=SD/4 is ≈ 1429. Considering a worst case scenario with *ε* = 0.3 for IgG percentage, we have ≈ 13333, 8333, and 3333 minimum antibody titer levels of stimulated neutralizing antibody. We thus predict that using even the pessimistic range of neutralizing antibody achieves the same level of high and medium-to-high neutralizing antibodies as observed in COVID-19 recovered individuals.

## 4 Discussion

In this work, we employ a mathematical model to study the vaccine-induced adaptive immune response through cell-mediated and humoral (antibody-mediated) immunity given an adenovirus vaccine. Using a set of nonlinear ordinary differential equations, we presented a new model of vaccine-induced immunity that was parameterized with the clinical trial data for the COVID-19 ChAdOx1-S (AZD1222) vaccine. The model parameters were determined by grid search over parameter ranges to minimize the RMSE of IFN*γ* and antibody functions. In addition to the fitted parameters, the model includes some chosen parameters. Our sensitivity analysis in Figure 3 shows that variations in these parameters do not significantly affect model peak values of the model parameters, except *mu*_21_, the activation rate of Th_0_ by the vaccine particles.

Our model predictions for IFN*γ* and antibody are consistent with the clinical trial data of the adenovirus-based Oxford vaccine [1]. Recent studies have explored scenarios for reduced vaccine-dose size, to consider if vaccine supply is scarce. In [40] the authors studied the effects of reducing the prime dose of a SARS-CoV2 adenovirus-based vaccine in a mouse model. Their in-vivo experiments demonstrated that mice initially primed with a low dose (LD) vaccine significantly exhibited a higher level of the immune response. In another study, Geoffroy et al. looked at the effects of increasing the time-interval between doses, using an SIR epidemiological model [41]. We have considered SD/LD cases with varying time-frames between doses. Our results show that an enhanced immune response can be realized in some immune response populations, whereas the antibody response is best if doses are given 28 days (4 weeks) apart.

Our mathematical model does not take into account memory B-cells and T-cells, instead focusing on correlations between such memory cells and the model peak plasma B- and Th_0_-cells. With more clinical data availability, future extension may include stimulation of memory cells. The inclusion of memory cells is a course for future work.

We have analyzed different scenarios given a second dose of vaccine that is delayed or reduced in size. Our model predictions show that either limiting the second dose or increasing the prime-boost time interval leads to an attenuated adaptive immune response. However, in agreement with previous clinical findings [3], the model predicted antibody achieves levels in the same range as the neutralizing antibody in 39% of COVID-19 recovered patients. In addition, with a minimum threshold for stimulated neutralizing antibody, the model shows similar immune protection to that of recovered patients. Hence, a delayed second dose in combination with smaller doses sizes may allow for sufficient dose allocation to meet specific population vaccination targets while maintaining vaccine efficacy.

It is important to note that [1] observed similar outcomes in LD/LD scenarios in their clinical trial compared to SD/SD individuals, across different age groups, given a 28-day interval between doses. We have not considered this case here, but plan to in future work.

## Data Availability

The data used in the manuscript is from the literature and is provided inside the paper.

## 5 Acknowledgments

We thank the Canadian In-Host Modelling Group (Matt Betti, Dan Coombs, David Dick, Thomas Hillen, Jude Kong, Kang Ling Liao, Nicole Mideo, Stephanie Portet, Angie Raad, Lindi Wahl, James Watmough) for fruitful discussions.

## 6 Author Contributions

Conceptualization, SF-S, HKO, JMH, MC; methodology, SF-S, HKO, JMH; software, SF-S, IRM, JMH; validation, SF-S, JMH, MC; formal analysis, SF-S, HKO, JMH; investigation, SF-S, JMH, MC, HKO, CK, SG; resources, SF-S, HKO, JMH, MSG; writing, SF-S, JMH; review and editing, SF-S, JMH, MC, IRM, CK, SG, MSG, HKO, JK; visualization, SF-S, JMH, MC, HKO; supervision, JMH; All authors have read and agreed to the published version of the manuscript.

## References

[1] Maheshi N Ramasamy, Angela M Minassian, Katie J Ewer, Amy L Flaxman, Pedro M Folegatti, Daniel R Owens, Merryn Voysey, Parvinder K Aley, Brian Angus, Gavin Babbage, et al. Safety and immunogenicity of chadox1 ncov-19 vaccine administered in a prime-boost regimen in young and old adults (cov002): a single-blind, randomised, controlled, phase 2/3 trial. The Lancet, 396(10267):1979–1993, 2020.

[2] Kylie M Quinn, Daniel E Zak, Andreia Costa, Ayako Yamamoto, Kathrin Kastenmuller, Brenna J Hill, Geoffrey M Lynn, Patricia A Darrah, Ross WB Lindsay, Lingshu Wang, et al. Antigen expression determines adenoviral vaccine potency independent of ifn and sting signaling. The Journal of clinical investigation, 125(3):1129–1146, 2015.

[3] Fan Wu, Aojie Wang, Mei Liu, Qimin Wang, Jun Chen, Shuai Xia, Yun Ling, Yuling Zhang, Jingna Xun, Lu Lu, et al. Neutralizing antibody responses to sars-cov-2 in a covid-19 recovered patient cohort and their implications. medRxiv, 2020.

[4] Lifang Zhang. Multi-epitope vaccines: a promising strategy against tumors and viral infections. Cellular & molecular immunology, 15(2):182–184, 2018.

[5] Tamalika Kar, Utkarsh Narsaria, Srijita Basak, Debashrito Deb, Filippo Castiglione, David M Mueller, and Anurag P Srivastava. A candidate multi-epitope vaccine against sars-cov-2. Scientific reports, 10(1):1–24, 2020.

[6] Ernesto Estrada. Covid-19 and sars-cov-2. modeling the present, looking at the future. Physics Reports, 2020.

[7] Charles A Janeway Jr, Paul Travers, Mark Walport, and Mark J Shlomchik. The complement system and innate immunity. In Immunobiology: The Immune System in Health and Disease. 5th edition. Garland Science, 2001.

[8] Angela S Clem. Fundamentals of vaccine immunology. Journal of global infectious diseases, 3(1):73, 2011.

[9] Jason R Lees. Interferon gamma in autoimmunity: A complicated player on a complex stage. Cytokine, 74(1):18–26, 2015.

[10] Víctor J Costela-Ruiz, Rebeca Illescas-Montes, Jose M Puerta-Puerta, Concepción Ruiz, and Lucia Melguizo-Rodríguez. Sars-cov-2 infection: The role of cytokines in covid-19 disease. Cytokine & growth factor reviews, 2020.

[11] Rhiannon Morris, Nadia J Kershaw, and Jeffrey J Babon. The molecular details of cytokine signaling via the jak/stat pathway. Protein Science, 27(12):1984–2009, 2018.

[12] Tadamitsu Kishimoto. Interleukin-6: discovery of a pleiotropic cytokine. Arthritis research & therapy, 8(2):1–6, 2006.

[13] Lauro Velazquez-Salinas, Antonio Verdugo-Rodriguez, Luis L Rodriguez, and Manuel V Borca. The role of interleukin 6 during viral infections. Frontiers in microbiology, 10:1057, 2019.

[14] Shizuo Akira and Tadamitsu Kishimoto. Il-6 and nf-il6 in acute-phase response and viral infection. Immunological reviews, 127:25–50, 1992.

[15] Toshio Tanaka, Masashi Narazaki, and Tadamitsu Kishimoto. Il-6 in inflammation, immunity, and disease. Cold Spring Harbor perspectives in biology, 6(10):a016295, 2014.

[16] JFA Po Miller and GF Mitchell. Cell to cell interaction in the immune response: I. hemolysin-forming cells in neonatally thymectomized mice reconstituted with thymus or thoracic duct lymphocytes. The Journal of experimental medicine, 128(4):801–820, 1968.

[17] Manfred Kopf, Heinz Baumann, Giulia Freer, Marina Freudenberg, Marinus Lamers, Tadamitsu Kishimoto, Rolf Zinkernagel, Horst Bluethmann, and Georges Köhler. Impaired immune and acute-phase responses in interleukin-6-deficient mice. Nature, 368(6469):339–342, 1994.

[18] Nanshan Chen, Min Zhou, Xuan Dong, Jieming Qu, Fengyun Gong, Yang Han, Yang Qiu, Jingli Wang, Ying Liu, Yuan Wei, et al. Epidemiological and clinical characteristics of 99 cases of 2019 novel coronavirus pneumonia in wuhan, china: a descriptive study. The lancet, 395(10223):507–513, 2020.

[19] Wang Wenjun, Liu Xiaoqing, Wu Sipei, Lie Puyi, Huang Liyan, Li Yimin, Cheng Linling, Chen Sibei, Nong Lingbo, Lin Yongping, et al. The definition and risks of cytokine release syndrome-like in 11 covid-19-infected pneumonia critically ill patients: Disease characteristics and retrospective analysis. MedRxiv, 2020.

[20] Puja Mehta, Daniel F McAuley, Michael Brown, Emilie Sanchez, Rachel S Tattersall, and Jessica J Manson. Covid-19: consider cytokine storm syndromes and immunosuppression. The lancet, 395(10229):1033–1034, 2020.

[21] L Chen, HG Liu, W Liu, J Liu, K Liu, J Shang, Y Deng, and S Wei. Analysis of clinical features of 29 patients with 2019 novel coronavirus pneumonia. Zhonghua jie he he hu xi za zhi= Zhonghua jiehe he huxi zazhi= Chinese journal of tuberculosis and respiratory diseases, 43:E005–E005, 2020.

[22] Hin Chu, Jasper Fuk-Woo Chan, Yixin Wang, Terrence Tsz-Tai Yuen, Yue Chai, Yuxin Hou, Huiping Shuai, Dong Yang, Bingjie Hu, Xiner Huang, et al. Comparative replication and immune activation profiles of sars-cov-2 and sars-cov in human lungs: an ex vivo study with implications for the pathogenesis of covid-19. Clinical Infectious Diseases, 71(6):1400–1409, 2020.

[23] Bo Diao, Chenhui Wang, Yingjun Tan, Xiewan Chen, Ying Liu, Lifen Ning, Li Chen, Min Li, Yueping Liu, Gang Wang, et al. Reduction and functional exhaustion of t cells in patients with coronavirus disease 2019 (covid-19). Frontiers in immunology, 11:827, 2020.

[24] Lan Dong, Jinhua Tian, Songming He, Chuchao Zhu, Jian Wang, Chen Liu, and Jing Yang. Possible vertical transmission of sars-cov-2 from an infected mother to her newborn. Jama, 323(18):1846–1848, 2020.

[25] Pan Luo, Yi Liu, Lin Qiu, Xiulan Liu, Dong Liu, and Juan Li. Tocilizumab treatment in covid-19: a single center experience. Journal of medical virology, 92(7):814–818, 2020.

[26] Jie Ma, Peng Xia, Yangzhong Zhou, Zhengyin Liu, Xiang Zhou, Jinglan Wang, Taisheng Li, Xiaowei Yan, Limeng Chen, Shuyang Zhang, et al. Potential effect of blood purification therapy in reducing cytokine storm as a late complication of critically ill covid-19. Clinical Immunology (Orlando, Fla.), 214:108408, 2020.

[27] Savannah F Pedersen, Ya-Chi Ho, et al. Sars-cov-2: a storm is raging. The Journal of clinical investigation, 130(5), 2020.

[28] Qiurong Ruan, Kun Yang, Wenxia Wang, Lingyu Jiang, and Jianxin Song. Clinical predictors of mortality due to covid-19 based on an analysis of data of 150 patients from wuhan, china. Intensive care medicine, 46(5):846–848, 2020.

[29] Dan Sun, Hui Li, Xiao-Xia Lu, Han Xiao, Jie Ren, Fu-Rong Zhang, and Zhi-Sheng Liu. Clinical features of severe pediatric patients with coronavirus disease 2019 in wuhan: a single center’s observational study. World Journal of Pediatrics, pages 1–9, 2020.

[30] Zhongliang Wang, Bohan Yang, Qianwen Li, Lu Wen, and Ruiguang Zhang. Clinical features of 69 cases with coronavirus disease 2019 in wuhan, china. Clinical infectious diseases, 71(15):769–777, 2020.

[31] Chaomin Wu, Xiaoyan Chen, Yanping Cai, Xing Zhou, Sha Xu, Hanping Huang, Li Zhang, Xia Zhou, Chunling Du, Yuye Zhang, et al. Risk factors associated with acute respiratory distress syndrome and death in patients with coronavirus disease 2019 pneumonia in wuhan, china. JAMA internal medicine, 180(7):934–943, 2020.

[32] Yang Yang, Chenguang Shen, Jinxiu Li, Jing Yuan, Minghui Yang, Fuxiang Wang, Guobao Li, Yanjie Li, Li Xing, Ling Peng, et al. Exuberant elevation of ip-10, mcp-3 and il-1ra during sars-cov-2 infection is associated with disease severity and fatal outcome. MedRxiv, 2020.

[33] Todd N. Eagar and Stephen D. Miller. 16 - helper t-cell subsets and control of the inflammatory response. In Robert R. Rich, Thomas A. Fleisher, William T. Shearer, Harry W. Schroeder, Anthony J. Frew, and Cornelia M. Weyand, editors, Clinical Immunology (Fifth Edition), pages 235–245.e1. Elsevier, London, fifth edition edition, 2019.

[34] Roald Nezlin. The immunoglobulins: structure and function. Academic Press, 1998.

[35] Harry W Schroeder Jr and Lisa Cavacini. Structure and function of immunoglobulins. Journal of Allergy and Clinical Immunology, 125(2):S41–S52, 2010.

[36] MD McKaya, RJ Beckmana, and WJ Conoverb. Comparison of three methods for selecting values of input variables in the analysis of output from a computer code. Technometrics, 21(2):239–245, 1979.

[37] Sally M Blower, Diana Hartel, Hadi Dowlatabadi, Robert M Anderson, and Roy M May. Drugs, sex and hiv: a mathematical model for new york city. Philosophical Transactions of the Royal Society of London. Series B: Biological Sciences, 331(1260):171–187, 1991.

[38] Jianyong Wu, Radhika Dhingra, Manoj Gambhir, and Justin V Remais. Sensitivity analysis of infectious disease models: methods, advances and their application. Journal of The Royal Society Interface, 10(86):20121018, 2013.

[39] Boloye Gomero. Latin hypercube sampling and partial rank correlation coefficient analysis applied to an optimal control problem. Thèse de doctorat. University of Tennessee, 2012.

[40] Sarah Sanchez, Nicole Palacio, Tanushree Dangi, Thomas Ciucci, and Pablo Penaloza-MacMaster. Limiting the priming dose of a sars cov-2 vaccine improves virus-specific immunity. bioRxiv, 2021.

[41] Félix Geoffroy, Arne Traulsen, and Hildegard Uecker. Vaccination strategies when vaccines are scarce: On conflicts between reducing the burden and avoiding the evolution of escape mutants. medRxiv, 2021.

